# Contrast Sensitivity Loss in Glaucoma Using the Reaction Time Paradigm

**DOI:** 10.1101/2025.10.10.25337590

**Authors:** Emmanuel Ezeuchu

## Abstract

Glaucoma is a progressive optic neuropathy characterized by retinal ganglion cell loss, often leading to visual impairment before measurable deficits in visual acuity occur. Contrast sensitivity (CS) provides critical insight into visual function, particularly under low-contrast conditions. This study investigates the pattern of contrast sensitivity loss in glaucoma using a reaction time (RT) paradigm, which reflects the interval between stimulus presentation and response initiation. Forty subjects (20 glaucoma patients, 20 healthy controls) aged 16 to 40 were tested using the Contrast Sensitivity Test Suite software with three stimuli (number, blinking square, jumping square). Reaction time increased as contrast decreased, with glaucoma patients showing significantly longer RTs across all stimuli compared to controls (p < 0.01). Regression analyses indicated that dysfunction in slow sustained (magnocellular) channels primarily accounts for low-contrast sensitivity deficits in glaucoma. These findings support the use of contrast sensitivity and reaction time measurements as sensitive functional markers for early glaucomatous changes, even in individuals with normal visual acuity.

## 1.0 INTRODUCTION

### Background

Glaucoma is a progressive optic neuropathy characterized by the loss of retinal ganglion cells, leading to irreversible visual field defects. Although visual acuity often remains normal in early stages, contrast sensitivity can be significantly impaired before any noticeable changes in vision occur. Contrast sensitivity refers to the ability to detect differences in luminance between an object and its background, and it plays a crucial role in everyday visual tasks such as reading, mobility, and facial recognition.

The integration of visual contrast stimuli is controlled by two primary channels: the fast transient (magnocellular, M) channels and the slow sustained (parvocellular, P) channels, which can be differentially affected in disease conditions (Shapley & Enroth-Cugell, 1973). Psychophysically derived measures, such as reaction time (RT) contrast sensitivity, provide insight into the functionality of these channels and have been shown to reflect the physiological contrast gain characteristics of neurons in the primary visual pathway (Kaplan & Shapley, 1986; Murray & Plainis, 2003; Sclar et al., 1990).

Reaction time paradigms have been used to reveal the contributions of these channels at different contrast levels. Fast transient channels dominate at high contrast, while slow sustained channels are more influential at low contrast. This makes RT paradigms particularly useful in detecting subtle functional deficits in glaucoma that may not yet be apparent through standard visual acuity tests.

#### Statement of the Problem

There are many clinical situations in which contrast sensitivity can be reduced while visual acuity remains normal. In healthy individuals, both visual acuity and contrast sensitivity remain stable until middle age, after which they decline gradually. In contrast, glaucoma can significantly reduce contrast sensitivity even before obvious ocular dysfunction manifests. This study investigates how contrast sensitivity and reaction time are affected in glaucoma patients compared with normal individuals.

### Aim of the Study

The aim of this study is to determine the pattern of contrast sensitivity loss in glaucoma using the reaction time paradigm.

### Objectives of the Study

1. To determine whether glaucoma has a significant effect on contrast sensitivity and reaction time.
2. To compare the effectiveness of the reaction time paradigm with the pulsed/steady-pedestal paradigm in detecting contrast sensitivity loss.

### Hypotheses

#### Null Hypotheses (Ho)

● Ho1: There is no significant pattern of contrast sensitivity loss in glaucoma using the reaction time paradigm.
● Ho2: Glaucoma has no significant effect on contrast sensitivity and reaction time.
● Ho3: There is no significant difference between the reaction time paradigm and the pulsed/steady-pedestal paradigm in detecting contrast sensitivity loss.

#### Alternate Hypotheses (HA)

● HA1: There is a significant pattern of contrast sensitivity loss in glaucoma using the reaction time paradigm.
● HA2: Glaucoma has a significant effect on contrast sensitivity and reaction time.
● HA3: There is a significant difference between the reaction time paradigm and the pulsed/steady-pedestal paradigm in detecting contrast sensitivity loss.

### Significance of the Study

This study will assist optometrists and ophthalmologists in identifying whether decreased contrast sensitivity in patients is associated with glaucoma. It highlights the importance of contrast sensitivity testing in suspected glaucoma cases and provides a foundation for future research linking contrast sensitivity with diagnostic measures for ocular diseases. Additionally, the findings may inform the development of more sophisticated contrast sensitivity and reaction time analyzers.

### Definition of Terms

● Contrast Gain: The increase in response per unit change in contrast.
● Pulse-Pedestal Paradigm: A spatio-temporal method for revealing parvocellular contrast gain.
● Steady-Pedestal Paradigm: A spatio-temporal method for revealing magnocellular contrast gain.
● Retino-Cortical Pathway: The visual pathway from the retina to the primary visual cortex, relaying visual information.

## 2. Methodology

### 2.1 Research Design

This prospective experimental study was conducted at Isoks Vision Eye Clinic, Asaba, Delta State, between 13th April and 20th July 2016.

#### 2.1.1 Sampling Method

Subjects were recruited using convenient stratified sampling.

#### 2.1.2 Sample Size

Forty subjects were included: 20 glaucoma patients and 20 healthy individuals. Sample size was estimated using the formula:

n=Z2⋅SD2E2n = \frac{Z^2 \cdot SD^2}{E^2}n=E2Z2⋅SD2

Where:

● nnn = sample size
● ZZZ = 1.96 for 95% confidence level
● SDSDSD = standard deviation
● EEE = absolute error

### 2.2 Study Population

Subjects aged 16–40 years with corrected visual acuity ≥0.47 logMAR were recruited. Both genders were included.

#### Inclusion criteria

● Glaucoma patients aged 16–40
● Healthy individuals aged 16–40
● Absence of ocular diseases such as cataract, diabetic or hypertensive retinopathy, optic neuritis, or macular degeneration

#### Exclusion criteria

● Age <16 or >40
● Visual acuity worse than 0.47 logMAR
● Muscular/connective tissue disorders affecting object handling

### 2.3 Materials

● Contrast Sensitivity Test Suite (Version 0.93)
● Computer (1366 × 768 resolution)
● Distance and near Snellen charts
● Ophthalmoscope
● Penlight
● Meter rule

### 2.4 Procedure

Subjects were examined to ensure eligibility. RT and CS were measured using three stimuli: number target, blinking square, and jumping square. Twenty-four contrast levels were presented per session. Each stimulus duration was 6000 ms, and subjects responded by clicking the target. RTs were recorded automatically by the software (Bailey et al., 2003).

### 2.5 Data Analysis

Independent Student’s t-tests were used to compare groups. SPSS version 22 was employed for statistical analysis.

### 2.6 Ethical Approval and Consent to Participate

Ethical approval for this study was obtained in 2016 from the following oversight bodies:

1. The Ethics Committee of the Department of Optometry at the University of Benin in Benin City, Nigeria, granted ethical approval for the study protocol.
2. The Ethics Committee of Isoks Vision Clinic in Asaba, Delta State, Nigeria, also granted ethical approval for the study protocol.

Written informed consent was obtained from all participants prior to enrollment. Participant confidentiality was strictly maintained, and all research procedures were conducted in full accordance with the ethical principles outlined in the Declaration of Helsinki (World Medical Association, 2013).

## 3.0 RESULTS

### 3.1 Participant Characteristics

A total of 40 subjects participated in this study, comprising 20 glaucoma patients and 20 healthy individuals with normal vision. The mean age for both groups was approximately 26.93 years, with both male and female participants represented. All subjects met the inclusion criteria, including corrected visual acuity of 0.47 logMAR or better.

### 3.2 Reaction Time Measurements

Reaction time measurements showed that the glaucoma group had a mean reaction time of 1636.82 ± 257.89 ms for the number stimulus, 1385.38 ± 188.38 ms for the blinking stimulus, and 1385.03 ± 173.16 ms for the jumping stimulus. In contrast, the normal (control) group had significantly faster reaction times, with a mean of 843.05 ± 120.38 ms, 777.11 ± 73.83 ms, and 764.59 ± 83.21 ms for the number, blinking, and jumping stimuli, respectively.

Statistical analysis revealed that these differences were significant (p = 0.00 for all stimuli), indicating that participants in the normal group responded more rapidly than those in the glaucoma group.

**Table 1.**
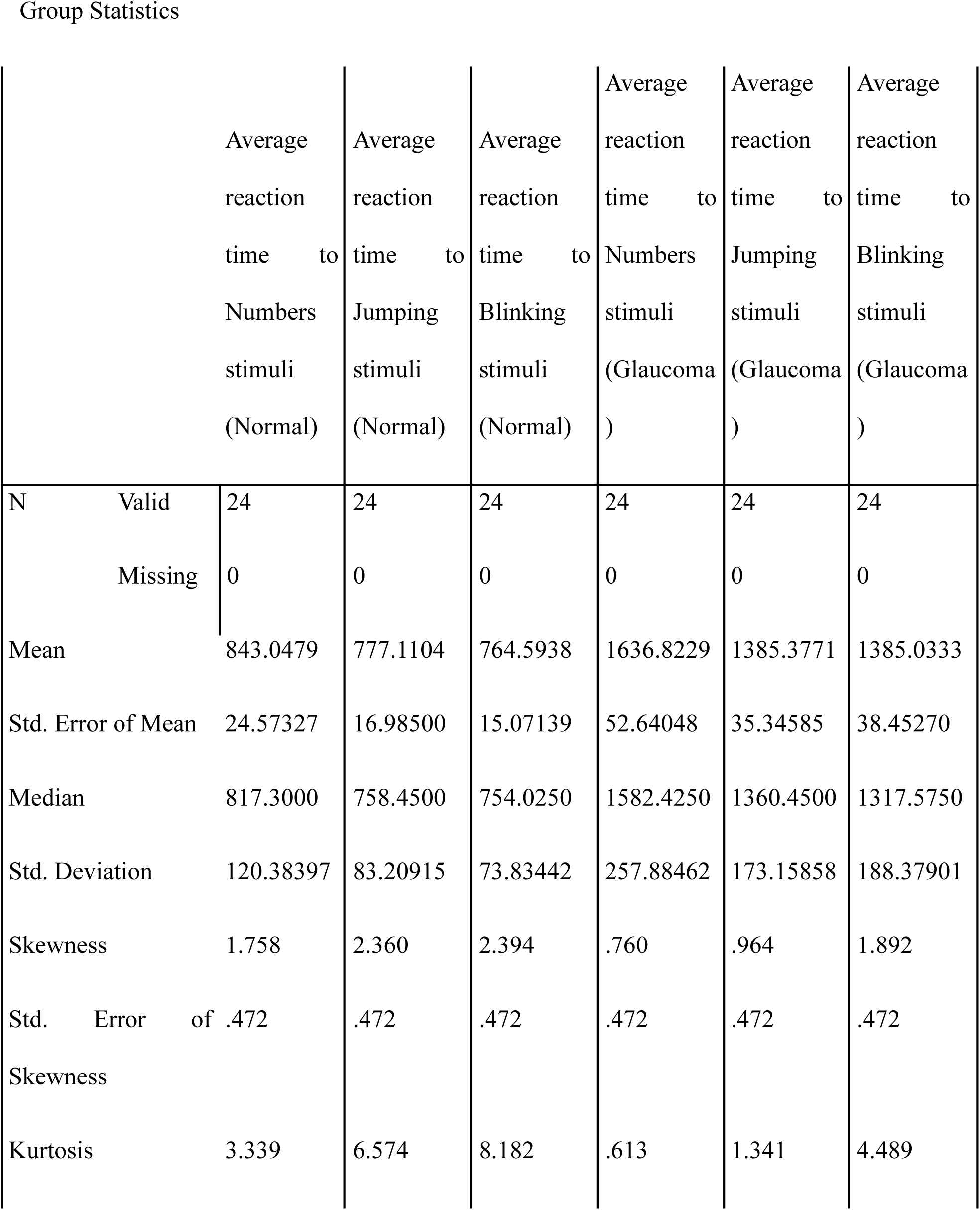
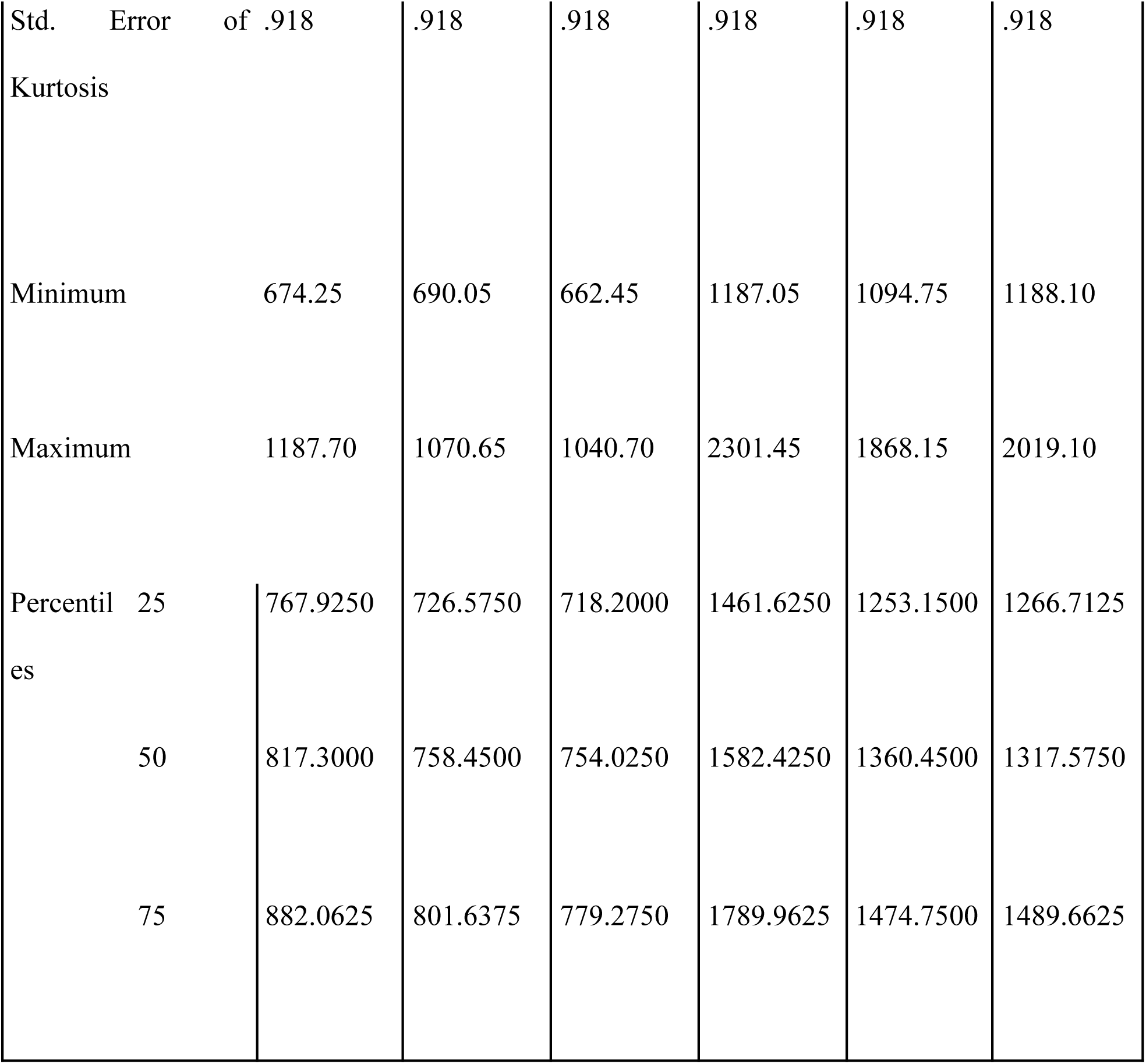
Mean, Median, Standard Error and Standard Deviation, of Reaction Time Between Glaucoma Group and Normal Group.

**Table 2:** Comparison of Reaction Time Result between the Glaucoma and Normal subjects. From the above table, (P = 0.000 i.e. P < 0.01) There is a significant difference in reaction time stimulus between the glaucoma individuals and normal individuals. The reaction times to the different stimuli used differ between Glaucoma individuals and normal individuals.

|  | Reaction time<br>to Number<br>stimulus | Reaction time<br>to Blinking<br>stimulus | Reaction time to Jumping stimulus |
| --- | --- | --- | --- |
| Mann-Whitney U | 1.000 | .000 | .000 |
| Wilcoxon W | 301.000 | 300.000 | 300.000 |
| Z | -5.918 | -5.938 | -5.938 |
| Asymp. Sig. (2-tailed) | .000 | .000 | .000 |
| Exact Sig. (2-tailed) | .000 | .000 | .000 |
| Exact Sig. (1-tailed) | .000 | .000 | .000 |
| Point Probability | .000 | .000 | .000 |

### 3.3 REGRESSION RESULTS FOR THE SUBJECTS

Plainis and Murray (2000) have described a wide range of stimulus characteristics in which the RT is a linear function of the reciprocal of contrast, so that

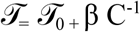

Where 𝒯 = reaction time, 𝒯_0_= the asymptotic reaction time, β = slope, C= contrast. Effectively this means the exponent in Pieron’s famous reaction time equation is -1. The advantage of using RTs is that sensitivity can be obtained over a range of supra-threshold contrasts and thereby provide a direct measure of contrast gain. Using a derivation of the well-known Naka -Rushton equation to obtain gains at high and low contrasts, Murray and Plainis (2003) have described a biphasic relationship, revealing the transition between two physiological mechanisms identified as reflecting M and P pathways.

**Table 3.**
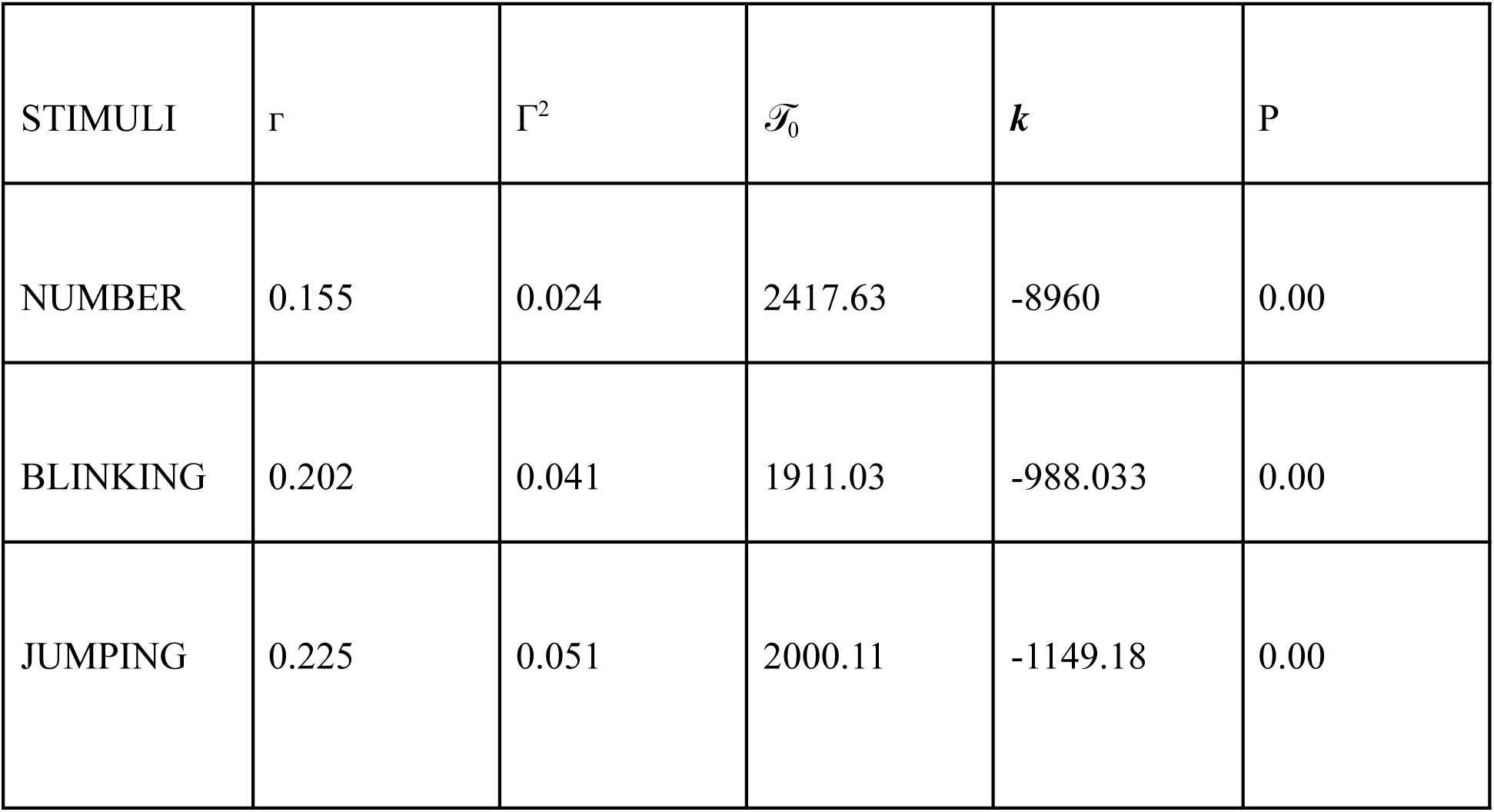
Model summary of the regression results for 20 Glaucoma patients.

**Table 4.** Model summary of the regression results for 20 Normal patients.

| STIMULI | $\Gamma$ | $\Gamma^2$ | $\mathcal{T}_0$ | $k$ | P |
| --- | --- | --- | --- | --- | --- |
| NUMBER | 0.156 | 0.24 | 816.63 | 131.88 | 0.01 |
| BLINKING | 0.38 | 0.001 | 770.22 | 23 | 0.404 |
| JUMPING | 0.004 | 0.00 | 765.89 | 2.157 | 0.936 |

### 4.0 DISCUSSION

The aim of this study was to determine the pattern of contrast sensitivity loss in glaucoma using the reaction time (RT) paradigm. This paradigm reveals the interval between the presentation of a visual stimulus and the initiation of a muscular response, serving as a useful tool to assess the rate of visual stimulus integration and interpretation. The integration of visual contrast stimuli has been shown to be controlled by two neural channels, the fast transient (magnocellular) and slow sustained (parvocellular) channels, which may be selectively affected in disease conditions (Shapley and Enroth-Cugell, 1973).

In this study, we employed a psychophysically derived measure of contrast gain, i.e., RT contrast sensitivity, to investigate the relative contribution of neural mechanisms at suprathreshold contrasts. This metric, which is reciprocal to the slope of the biphasic RT versus C⁻¹ function, has been shown to resemble the physiological contrast gain used to model neuronal contrast response characteristics in the primary visual pathway (Kaplan and Shapley, 1986; Murray and Plainis, 2003; Sclar et al., 1990).

The contrast sensitivity and reaction time functions revealed the transition from the operation of fast transient channels (at high contrast levels) to slow sustained channels (at low contrast levels). These two mechanisms largely determine the variation of RT with contrast sensitivity (CS). Accordingly, the discussion of our findings focuses on the pattern of contrast sensitivity loss, the zones of influence for each channel, and the slope that reflects how CS loss affects RT.

The graphs obtained from our results demonstrate a distinct difference in the pattern of contrast sensitivity loss between the two groups. The difference in mean reaction times between glaucoma and normal subjects was statistically significant (p < 0.01), supporting the alternate hypothesis (HA1) and rejecting the null hypothesis (H0_1_). A decrease in contrast sensitivity corresponded with an increase in reaction time.

The contrast sensitivity curves for glaucoma subjects (Fig. 1) were steeper at low contrast levels compared to those of normal subjects (Fig. 2). This finding agrees with the slope of the RT-CS function for each subject in this range. It supports the observations of Murray and Plainis (2003), who proposed that at low spatial frequencies, the reciprocal contrast function exhibits shallow slopes reflecting high contrast gain associated with magnocellular (M) processing, whereas at higher spatial frequencies, steeper slopes reflect lower gain typical of parvocellular (P) processing.

**Fig 1.**
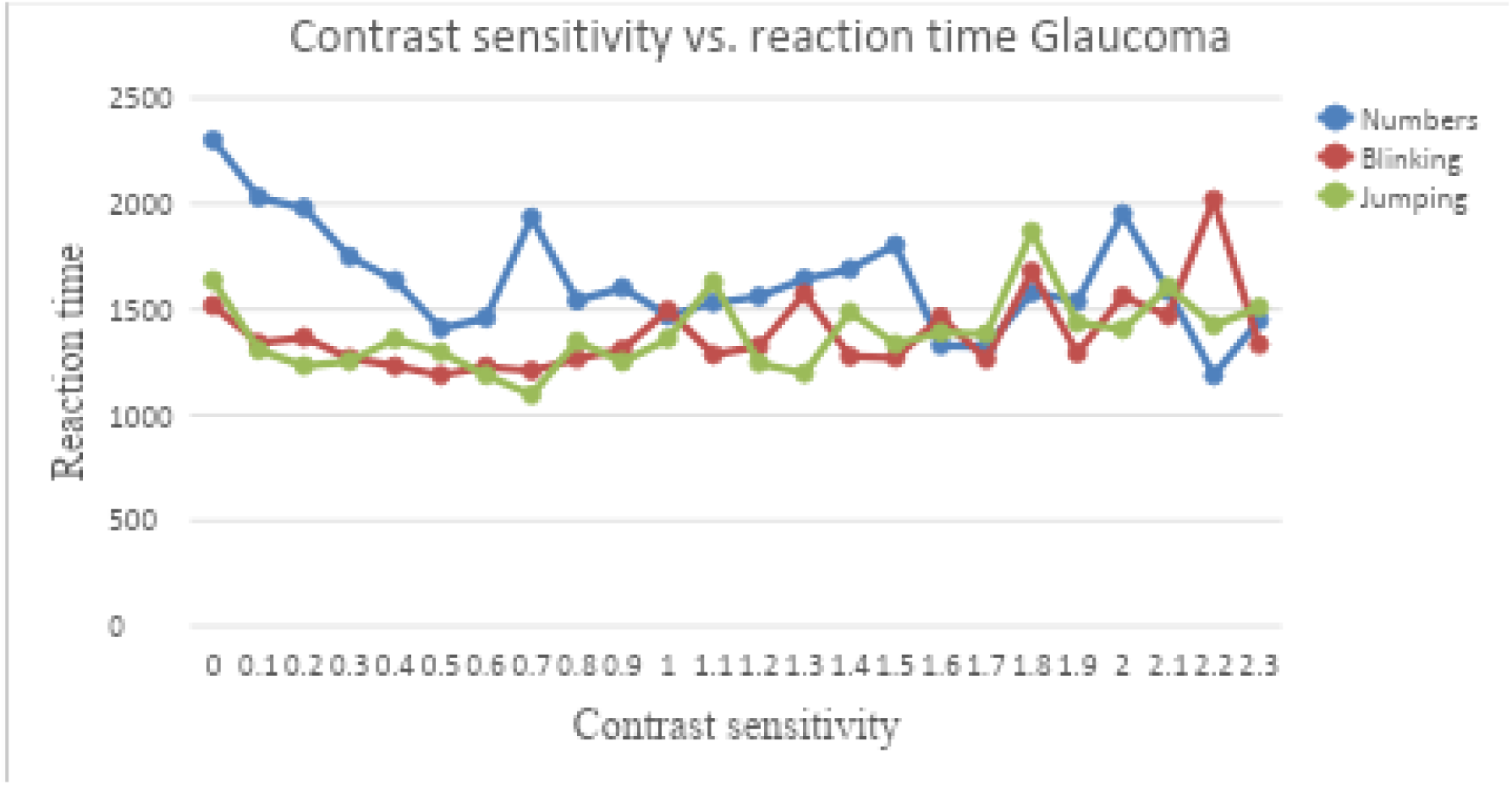
Graph showing the Contrast Sensitivity and Reaction Time for Glaucoma group. Interpretation: From the graph above, we can see the general pattern of contrast sensitivity and reaction time curve in the glaucoma group. The reaction is very high and increases as the contrast level decreases. There is an irregular flow which was assumed to start at a contrast level of 0.6log unit this reveals the declining response from the slow sustained channels (MC channels)

**Fig 2.**
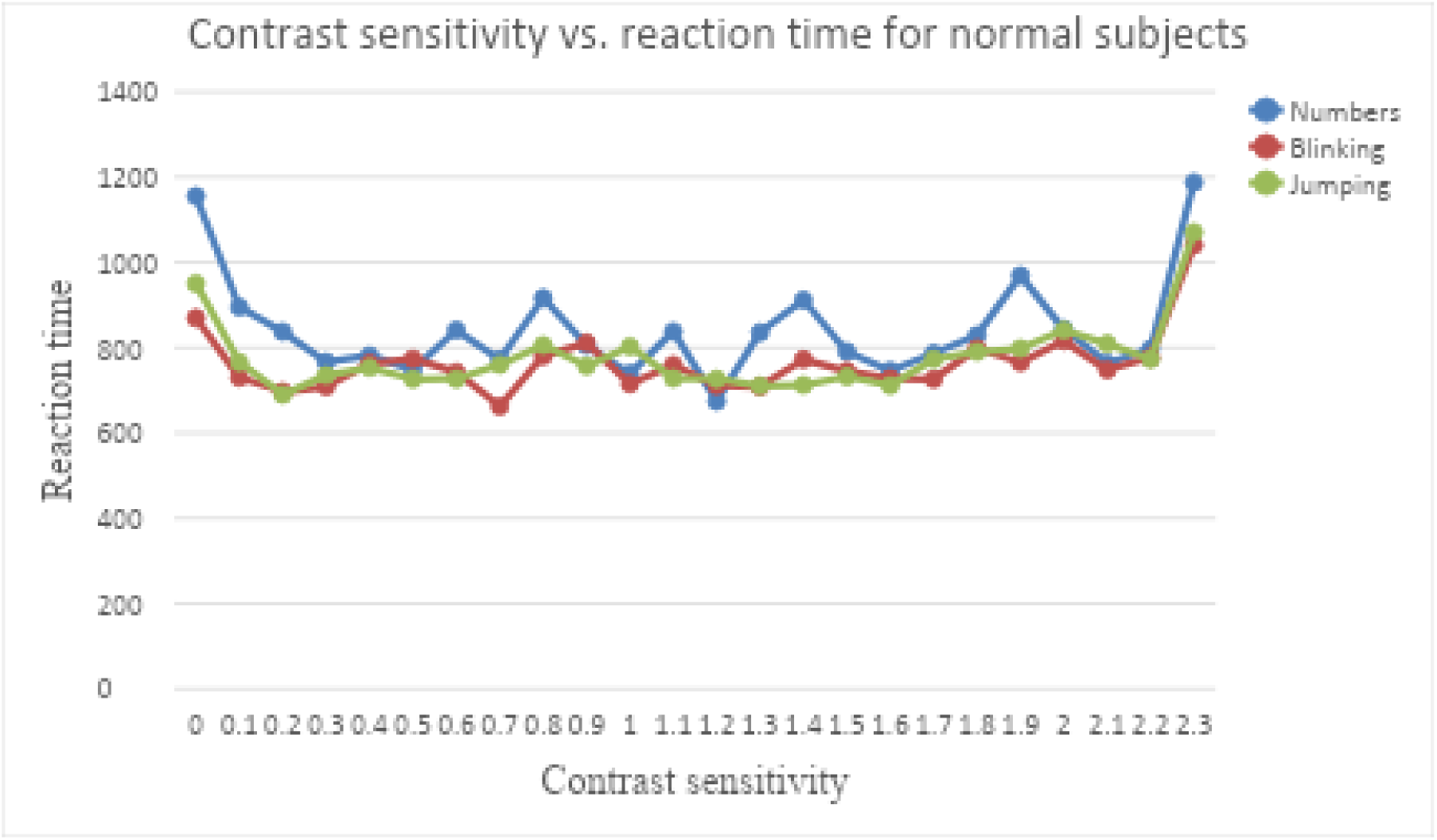
Graph showing the Contrast Sensitivity and Reaction Time for Normal group. Interpretation: From the graph above, we can see the general pattern of contrast sensitivity and reaction time curve for normal group. Reaction time in this graph is low and maintains a steady flow until it reaches the contrast level of 2.2log unit. This steady flow suggests a normal activity of the slow sustained channels (MC channel)

**Fig 3.**
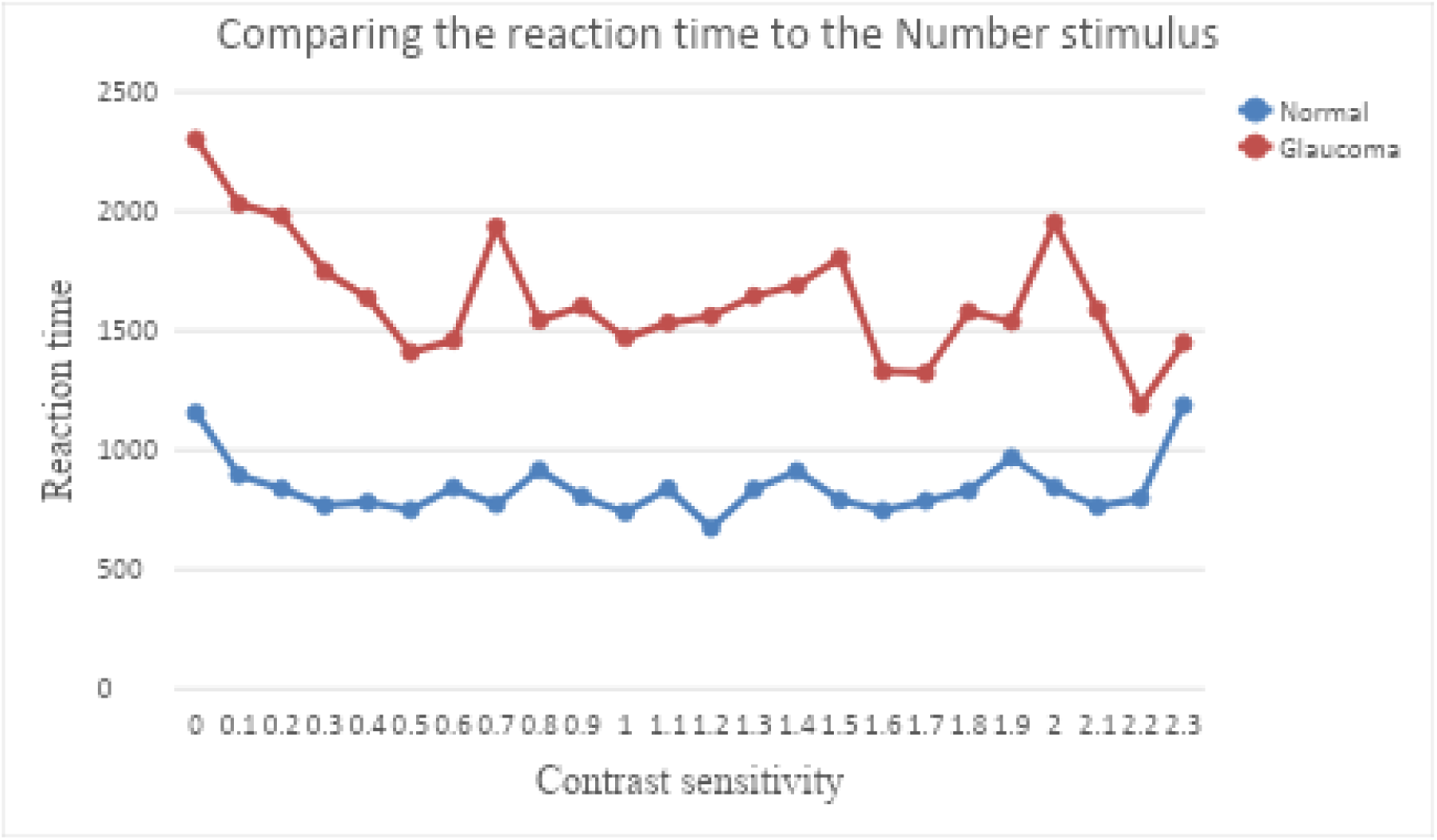
Graph comparing the Contrast Sensitivity and Reaction Time for the Number stimulus for glaucoma and Normal group. Interpretation: From the graph above, we can see that there is a difference in contrast sensitivity and reaction time for the glaucoma group and normal group for the number stimulus.

**Fig 4.**
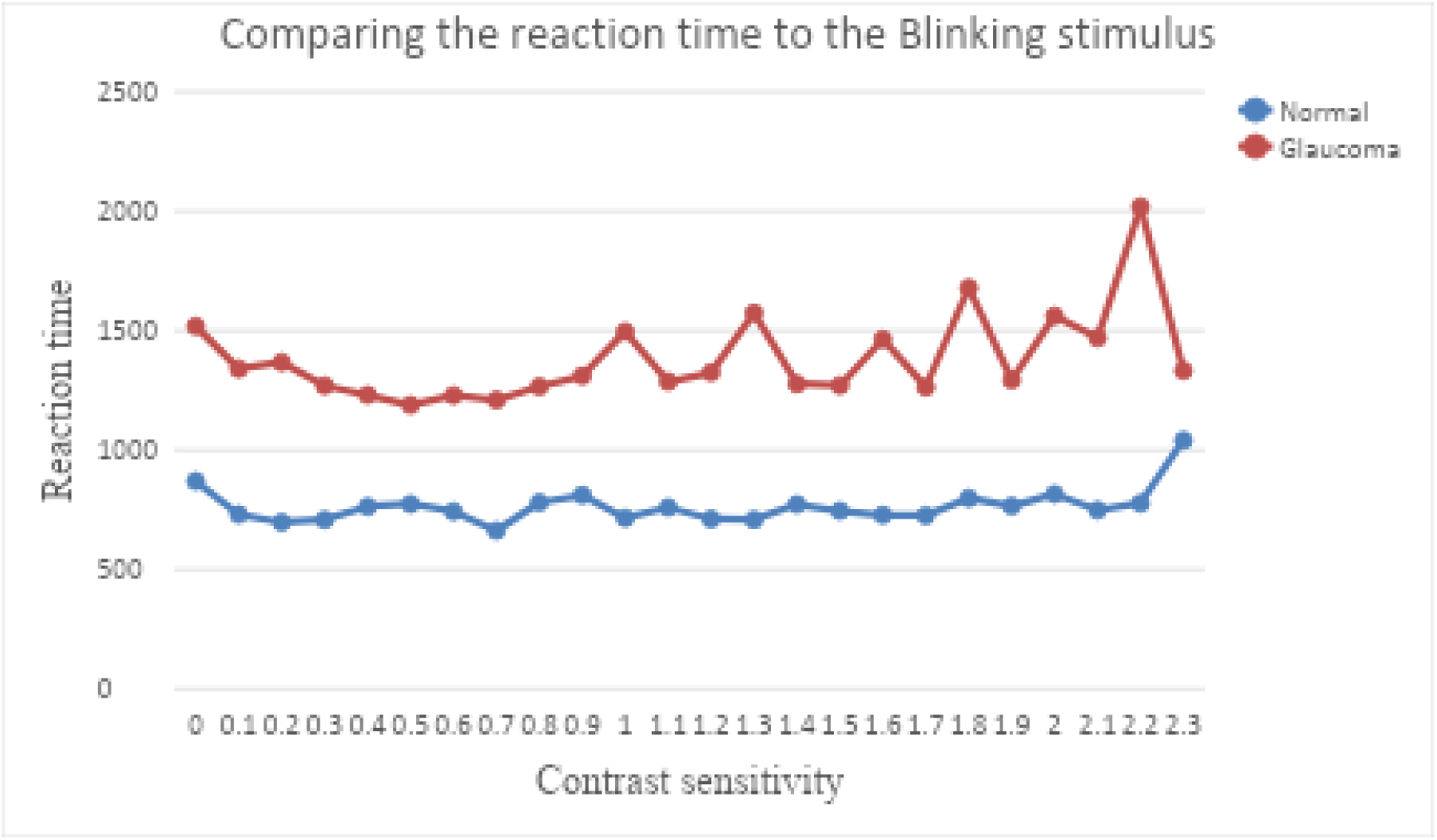
Graph comparing the Contrast Sensitivity and Reaction Time for the blinking stimulus for glaucoma and Normal group. Interpretation: From the graph above, we can see that there is a difference in contrast sensitivity and reaction time for the glaucoma group and normal group for the blinking stimulus.

**Fig 5.**
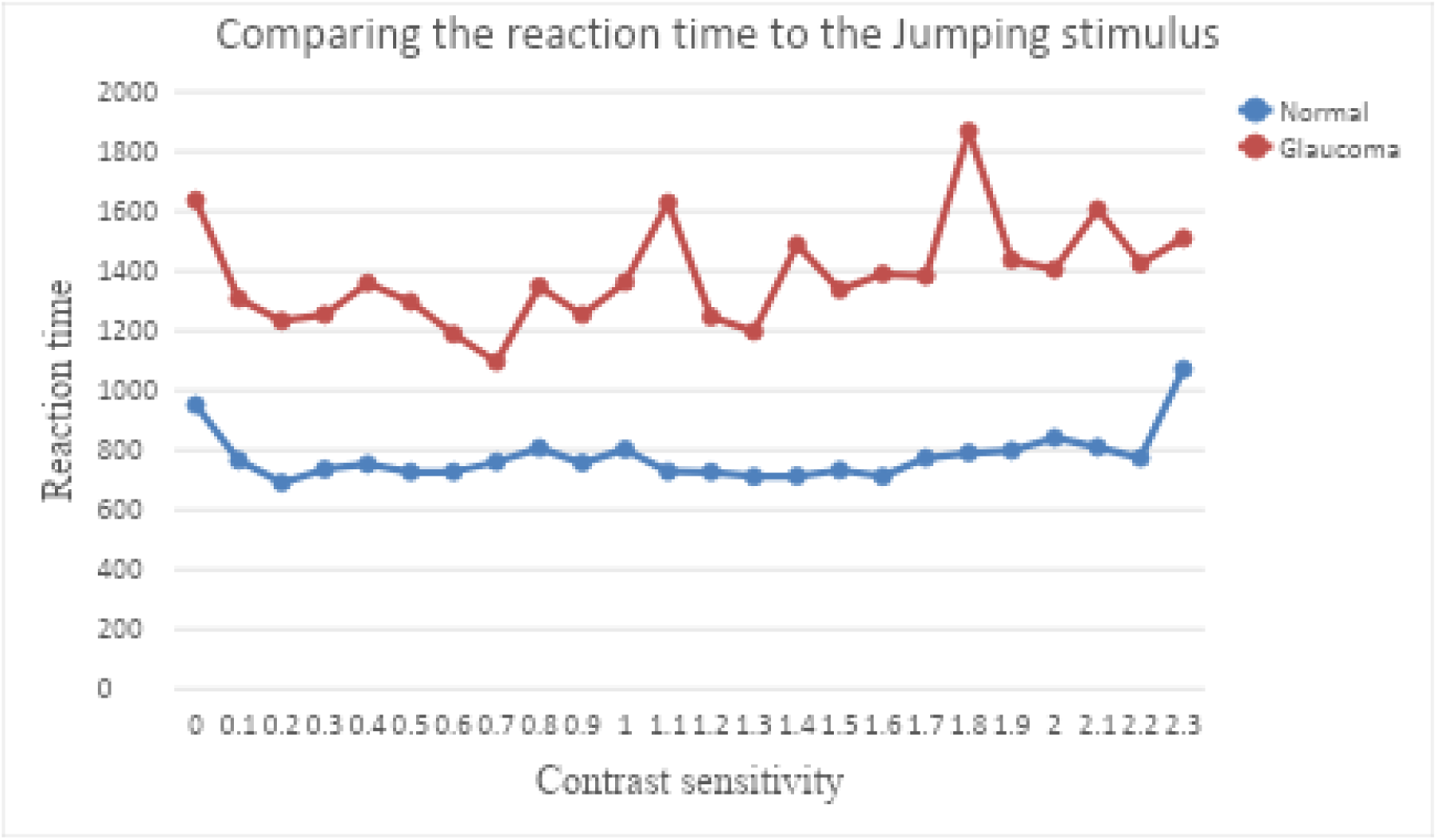
Graph comparing the Contrast Sensitivity and Reaction Time for the jumping stimulus for glaucoma and Normal group. Interpretation: From the graph above, we can see that there is a difference in contrast sensitivity and reaction time for the glaucoma group and normal group for the jumping stimulus.

Reaction time increased as contrast level decreased, with the glaucoma group showing a faster rate of increase due to reduced contrast sensitivity. This suggests dysfunction in the slow sustained (magnocellular) channels. This observation is consistent with previous findings (Murray and Plainis, 2003), which reported that a decrease in contrast sensitivity corresponds to an increase in reaction time.

An important objective of this research was to ascertain the effect of glaucoma on contrast sensitivity and reaction time. The data clearly demonstrated a significant effect, leading to acceptance of the alternate hypothesis (HA2) and rejection of the null hypothesis (H0_2_).

A second objective was to compare the reaction time paradigm with the pulsed/steady-pedestal paradigm in detecting contrast sensitivity loss. Our findings were congruent, revealing that reaction time increases as contrast sensitivity decreases, consistent with previous reports using the pulsed/steady-pedestal paradigm. Therefore, the alternate hypothesis (HA3) was accepted, and the null hypothesis (H0_3_) was rejected.

The negative slope value in the regression analysis revealed a rapid decrease in reaction time as contrast level increased. This simple linear relationship suggests that either a single mechanism M or P, depending on stimulus conditions, operates across the entire contrast range. In this study, low contrast levels were predominantly affected, supporting the notion that dysfunction of the magnocellular system (slow sustained channel) is primarily responsible for contrast sensitivity loss in glaucoma.

Glaucoma is no longer viewed solely as a disease of elevated intraocular pressure and optic cup changes. Studies have shown that more than 10% of retinal ganglion cells may be lost before a measurable decline in visual acuity is observed. However, long before this stage, a significant decline in contrast sensitivity occurs. This underscores the importance of assessing contrast sensitivity and reaction time as potential early indicators of glaucomatous visual dysfunction.

### 5.0 CONCLUSION

The results of this study suggest that contrast sensitivity loss in glaucoma follows a predictable pattern. Using the reaction time paradigm, an approach designed to assess contrast discrimination channels, we found that the channels responsible for detecting low contrasts, primarily the magnocellular pathways, were most affected in glaucoma. This disruption forms the basis for the reduced contrast sensitivity commonly observed in glaucomatous vision.

Our findings reinforce established neurophysiological principles: when both magnocellular (M) and parvocellular (P) systems are engaged, the M system predominates near-threshold reaction times, while the P system dominates at higher contrasts due to the saturation of the M system.

Importantly, the study demonstrates that even in individuals with good visual acuity, glaucoma can produce measurable declines in functional vision. The observed increase in reaction time reflects this subtle impairment, highlighting the potential of reaction time paradigms as a sensitive tool for detecting early glaucomatous changes in visual processing.

### 6.0 RECOMMENDATIONS

It is strongly advised that contrast sensitivity testing be included as a routine part of eye examinations. This will help in detecting early functional changes in retinal cells responsible for contrast discrimination, especially before noticeable loss in visual acuity occurs.

Contrast sensitivity should be closely monitored in all patients diagnosed with, or at risk of, glaucoma. In addition, contrast enhancement strategies should be considered during the management and rehabilitation of glaucoma patients to improve their functional vision and quality of life.

Optometrists, ophthalmologists, and other eye care professionals should be aware that contrast sensitivity can decrease significantly in glaucoma even when visual acuity remains normal. This awareness should guide comprehensive patient evaluation and timely intervention.

## Data Availability

All data produced in the present study are available upon reasonable request to the authors

## ACKNOWLEDGEMENTS

The author sincerely appreciates the supervision and guidance of Dr. G. A. Akinlabi of the Department of Optometry, University of Benin, Benin City, Nigeria, whose mentorship was invaluable throughout the course of this research.

Special thanks go to the management and staff of Isoks Vision Eye Clinic, Asaba, Delta State, for providing the facilities and support necessary to carry out this study.

The author also expresses heartfelt gratitude to all study participants for their time, patience, and cooperation.

